# Reference interval of optimal vitamin D level for an adult population of Bangladesh

**DOI:** 10.1101/2024.11.07.24316898

**Authors:** Wasim Md Mohosin Ul Haque, Jalaluddin Ashraful Haq, Md. Faruque Pathan, Mohammed Abu Sayeed

**Affiliations:** Department of Nephrology, Bangladesh Institute of Research and Rehabilitation for Diabetes Endocrine and Metabolic Disorders, Dhaka, Bangladesh; Department of Microbiology, Ibrahim Medical College, Dhaka, Bangladesh; Department of Endocrinology, Bangladesh Institute of Research and Rehabilitation for Diabetes Endocrine and Metabolic Disorders, Dhaka, Bangladesh; Department of Community Medicine, Ibrahim Medical College, Dhaka, Bangladesh

**Keywords:** Vitamin D, parathyroid hormone, reference interval, cut-off value, deflection point, coastal fishermen, urban population, Bangladesh

## Abstract

Vitamin D deficiency presents a significant public health concern, especially in regions where reference intervals from Western populations may not apply due to differences in sun exposure and ethnicity. This study aimed to establish population-specific reference intervals for serum 25-hydroxyvitamin D [25(OH)D] and to determine a deficiency cutoff for healthy adults in Bangladesh. In a cross-sectional design, we assessed serum 25(OH)D and intact parathyroid hormone (iPTH) levels in 125 coastal fishermen (Group 1) and 371 urban residents (Group 2), comprising healthy adults aged 18 years or older. Group 1 served as a reference to establish baseline vitamin D levels, while Group 2 data aided in determining the deficiency cutoff. Measurements were conducted using chemiluminescent immunoassay, and reference intervals were calculated according to Clinical and Laboratory Standards Institute (CLSI) Guidelines C28-A3. The deficiency cutoff was identified at the deflection point of iPTH levels. Results indicate a reference interval for serum 25(OH)D of 15.88–45.27 ng/ml among coastal fishermen. Among urban residents, mean serum 25(OH)D was 21.53 ± 15.98 ng/ml, with iPTH levels showing significant increases below 12.16 ng/ml (95% CI: 11.04–13.28), establishing this as the deficiency cutoff. Urban residents exhibited significantly lower vitamin D levels than coastal fishermen (21.53 ng/ml vs. 27.36 ng/ml, p < 0.001). Limitations include potential selection bias due to convenience sampling and the use of chemiluminescent immunoassay instead of the gold-standard LC-MS/MS assay. This study provides the first population-specific reference intervals for serum 25(OH)D in Bangladesh, accounting for unique sun exposure patterns and ethnic factors, and sets a deficiency threshold at 12.16 ng/ml. These findings are critical for guiding targeted interventions against vitamin D deficiency in this region.

## Introduction

Vitamin D, a crucial fat-soluble vitamin, plays a vital role in various physiological functions, including calcium and phosphorus homeostasis, immune function, and cell growth modulation(1–4). Consequently, its deficiency can lead to a range of diseases and disabilities (3, 5, 6). Despite Bangladesh’s favourable geographical location, the prevalence of vitamin D deficiency remains high even among seemingly healthy individuals (7, 8).

Current reference intervals for vitamin D are predominantly derived from studies conducted in Western populations and may not accurately reflect the physiological needs or health outcomes of diverse global populations(9, 10). Variations in genetics, skin pigmentation, dietary habits, and sun exposure significantly impact vitamin D levels, rendering universal reference intervals potentially inappropriate for many non-Western populations. Genetic polymorphisms affecting vitamin D metabolism and receptor activity vary among ethnic groups, leading to different optimal levels of serum 25-hydroxyvitamin D [25(OH)D] required for health (11, 12). Consequently, a one-size-fits-all approach can result in incorrect diagnoses, unnecessary supplementation, potential toxicity, or under-treatment and persistence of deficiency-related health issues.

Therefore, establishing a vitamin D reference interval specific to Bangladesh is crucial for accurate diagnosis and effective management of vitamin D deficiency. This study aims to establish the reference interval of serum 25-hydroxyvitamin D [25(OH)D] levels for the adult population of Bangladesh by examining two distinct groups: adequately sun-exposed healthy coastal fishermen and urban individuals with limited sun exposure.

Also, the study determined the lower cut-off value of vitamin D deficiency level in Bangladeshi adults by detecting the deflection point of serum intact parathyroid hormone (iPTH) level compared to serum 25(OH)D level. Vitamin D deficiency physiologically induces increased synthesis/production of parathyroid hormone.

## Methodology

The study was conducted from January 1, 2017, to December 31, 2021. The study protocol was approved by the institutional ethics review board. Informed consent was obtained from all participants, and data confidentiality was strictly maintained.

### Study site and population

The study was conducted in two geographical locations: Cox’s Bazar, a coastal district, about 298 km south of Dhaka city along the coast of Bay of Bengal with abundant sunlight; and Dhaka, the capital city, characterized by limited sun exposure of its inhabitants due to urbanization. The study was conducted from January 1, 2017, to December 31, 2021.

### Study design

Cross-sectional study design was employed to assess vitamin D levels in two population groups: adequately sun-exposed healthy coastal fishermen (Group-1) and urban residents with limited sun exposure (Group-2). Group-1 consisted of healthy adult coastal fishermen aged 18 years and above from Cox’s Bazar, who are regularly and adequately exposed to sunlight and consume marine fish, a rich source of vitamin D. Adequate sun exposure was defined as exposure to direct sunlight at least for 30 minutes, between 11 am and 2 pm, three times a week (13). Group-1 individuals (coastal fishermen) were considered as reference population for determining the vitamin D reference interval of local population. Group-2 comprised of ostensibly healthy urban residents from Dhaka, aged 18 years and above, with limited sun exposure due to lifestyle and environmental factors.

### Exclusion Criteria

Any individual who refused to participate in the study or who were physically and psychologically handicapped, had known acute or chronic illnesses like diabetes mellitus, hypertension, liver disease or gastrointestinal disorders, kidney disease, malignancy, metabolic bone disease, primary hyperparathyroidism or lactose intolerance were excluded. Anyone with relevant biochemical abnormalities and on anti-epileptic drugs was also excluded.

For Group-1, anyone who was consuming either vitamin D or calcium-containing products or who had ever consumed such drugs in the past was excluded.

### Sampling technique and sample size

Convenience sampling was used to select participants from both groups. The sample size was determined based on the desired confidence limits and the variability in vitamin D levels observed in preliminary studies. Adequate sample sizes were ensured to provide statistically significant results.

### Data collection tools and procedures

Sociodemographic and clinical data were collected using structured questionnaires. Height, weight, and blood pressure were measured, and blood samples were collected for estimation of serum 25(OH)D, iPTH levels and relevant biochemical parameters. The biochemical analyses were conducted using standardized procedures to ensure accuracy and reliability.

### Collection of blood and biochemical investigations

About 10 ml ofvenous blood was collected aseptically from each participant after counselling and thorough clinical evaluation. Serum was separated immediately and was preserved at –60^0^C for biochemical analysis. Chemiluminescent micro-particle immunoassay (CMIA) was used to estimate serum 25(OH)D level (14) by Architect analyser. Based on the Endocrine Society guideline (9), **v**itamin D level was categorised as: adequate/normal range: 75-100 nmol/L (30-40ng/ml), insufficient: 50 – 74.9 nmol/L (20ng/ml-29.99ng/ml) and deficient: <50 nmol/L (<20ng/ml). Samples were tested for random blood glucose (RBG), serum creatinine, calcium, PO4, albumin, alkaline phosphatase and iPTH by Architect autoanalyser at the biochemistry laboratory of BIRDEM.

### Statistical Analyses

Statistical analyses were conducted using descriptive and inferential methods to summarize the data and assess factors influencing vitamin D levels. Chi-square tests, correlation analyses, and regression models were employed to explore these relationships. MedCalc version 14 statistical software was utilized to calculate reference intervals (95%, double-sided) following parametric, non-parametric, and robust methods as outlined in the CLSI C28-A3 guideline(15). A 90% confidence interval (CI) was used for these reference intervals, applying the appropriate statistical techniques.

To determine the lower cutoff value of vitamin D deficiency, the deflection point of serum iPTH levels was identified using a non-linear model. Piecewise linear regression, applied through the Python plugin, was used to fit two separate lines before and after the deflection point (or “break” point), where the relationship between vitamin D and PTH shifts.

The equation used in this case represented as follows:

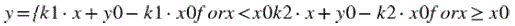

In this model:

- x represents Vitamin D level (independent variable).
- y represents PTH level (dependent variable).
- x_0_ is the deflection point where the two lines meet.
- y_0_ is the PTH level at the deflection point.
- k_1_ is the slope of the line before the deflection point (for x<x_0_).
- K_2_is the slope of the line after the deflection point (for x≥x_0_).

The steps for calculating the deflection point include:

1. **Initial estimation:** A starting guess for x_0_ is made based on visual inspection or prior knowledge.
2. **Curve fitting:** The SciPy curve fit function optimizes x_0_, y_0_, k_1_and k_2_ by minimizing the difference between the model and observed data.
3. **Confidence intervals:** Standard errors are used to calculate 95% confidence intervals for the estimates.

This approach effectively identifies the point where the relationship between vitamin D and PTH levels changes.

### Ethical Consideration

The Ethics Review Committee (ERC) of the Centre for Higher Studies and Research, Bangladesh University of Professionals and Diabetic Association of Bangladesh has approved the protocol. Written informed consent was obtained from all participants prior to study enrolment and blood sample collection. The aims and objectives of the study, as well as its procedure, were explained to each participant. Participants were assured that all information and records would be kept confidential and only used for research purposes. Participants had the right to withdraw from the study at any time.

## Result

Total 125 and 371 individuals were enrolled in Group 1 and Group 2 respectively. Group-1 study population had a mean age of 37.98±11.61 years, (95% CI :35.93, 40.04), and a mean BMI of 22.22±3.15 kg/m², (95% CI:21.66, 22.77). Detail anthropometric and biochemical parameters of Group-1 study population is shown in Table 1.

**Table 1:** Anthropometric and biochemical parameters of Group-1(coastal fishermen) study population (n=125)

| Parameters | n(%) | Mean±SD | 95% CI of Mean |
| --- | --- | --- | --- |
| Age(years) |  |  |  |
| 18-30 | 40(32.0) | 25.48±3.68 | 24.3, 26.65 |
| 31-40 | 40(32.0) | 35.90±2.97 | 34.95, 36.85 |
| 41-50 | 28(22.4) | 46.68±3.32 | 45.39, 47.97 |
| >50 | 17(13.6) | 58.00±3.35 | 56.28, 59.72 |
| Total | 125 | 37.98±11.61 | 35.93, 40.04 |
| Height(cm) | 125 | 161.10±7.57 | 159.76, 162.44 |
| Weight(kg) | 125 | 57.52±8.37 | 56.04, 59.00 |
| BMI |  |  |  |
| <18.5 | 10(8.0) | 16.69±1.69 | 15.48, 17.90 |
| 18.5-24.9 | 91(72.8) | 21.57±1.74 | 21.21, 21.93 |
| 25-29.9 | 24(19.2) | 26.97±1.46 | 26.35, 27.58 |
| RBS (mmol/l) | | 6.01±0.87 | 5.85, 6.16 |
| S creatinine (mg/dl) | | .8992±0.11 | 0.88, 0.92 |
| S albumin (g/L) | | 45.4024±2.92 | 44.89, 45.92 |
| S calcium (mg/dl) | | 9.3720±0.44 | 9.29, 9.45 |
| S PO4 (mg/dl) | | 3.2888±0.54 | 3.19, 3.38 |
| S Alk Phos (U/L) | | 79.86±16.38 | 76.96, 82.76 |
| S iPTH (pg/ml) | | 20.6264±8.85 | 19.06, 22.19 |

Note: RBS= random blood sugar, Alk Phos= alkaline phosphatase (Normal:44 to 147 IU/L), iPTH= intact parathyroid hormone. (Normal:10-65 pg/ml)

Group-2 included a total of 371 urban healthy adults. Among the 371 participants, 47.2% were male and 52.8% were female, with a mean age of 44.19 ± 11.48 years. BMI of the majority of the participants were between 18.5 to 29.9 kg/m²). Detail socio-demographic and anthropometric characteristics of Group-2 study population is shown in Table 2.

**Table 2.**
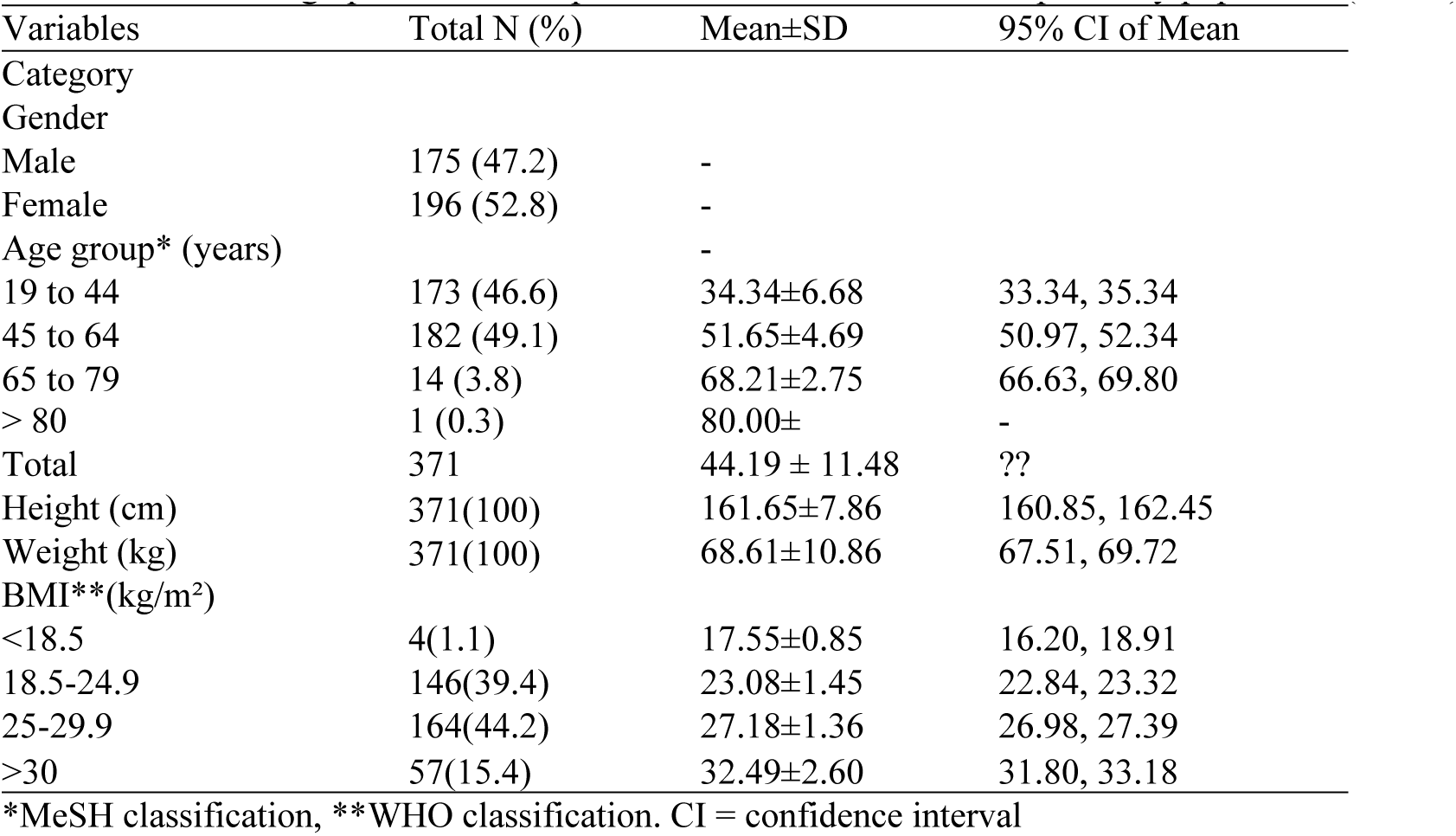
Socio-demographic and anthropometric characteristics of Group-2 study population.

The mean serum vitamin D level of Group 1 study population was 27.36±7.27 ng/ml, (95% CI:26.08, 28.65). No significant difference was found in vitamin D levels amongst age groups. However, there was a significant difference among BMI groups (F = 3.377, p = .037). Post-hoc testing using Bonferroni correction revealed that individuals with the highest BMI (25-29.9 kg/m²) had significantly (p = .037) lower serum 25(OH)D levels compared to those with the lowest BMI (<18.5 kg/m²), with a mean difference of –6.82 ng/ml. No significant differences were observed between other BMI groups (Table 3).

**Table 3.**
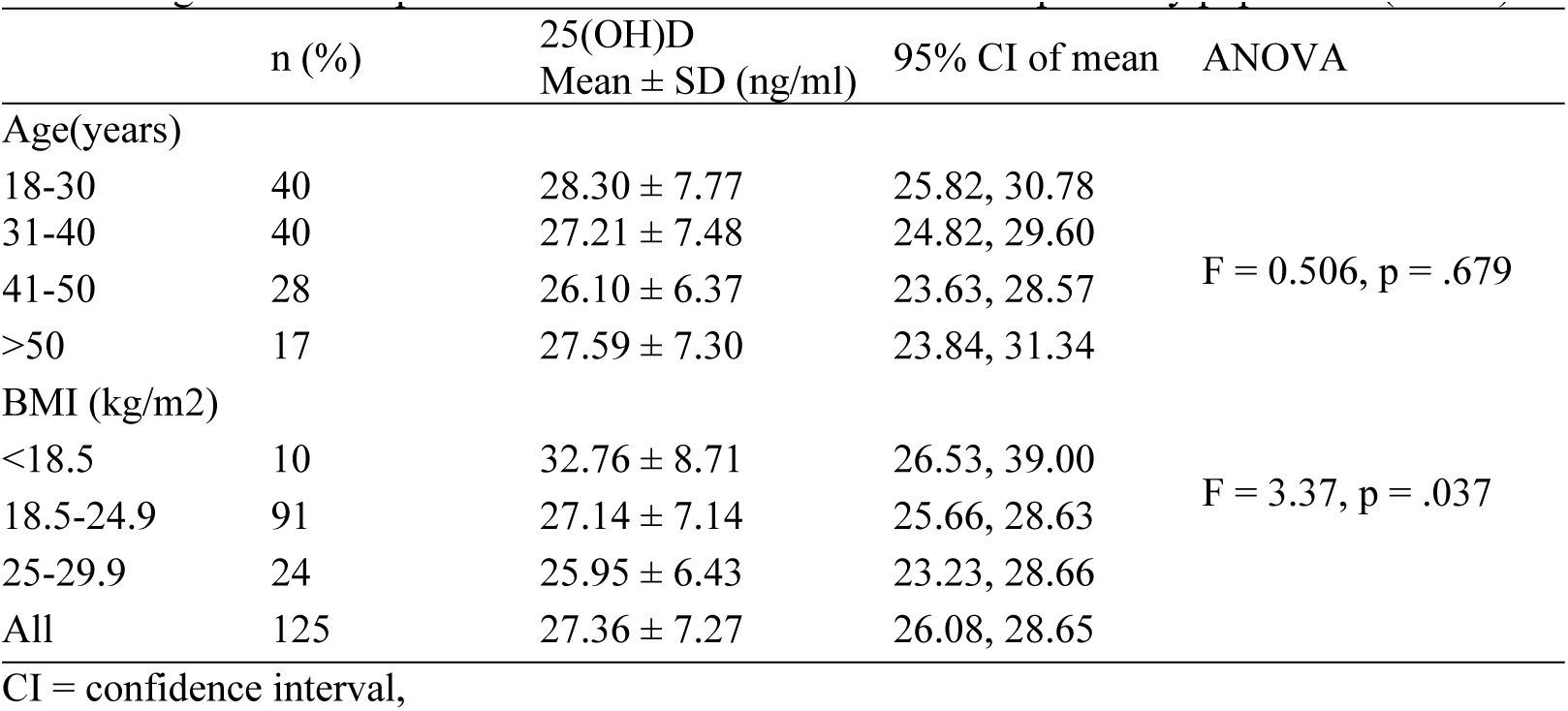
Age and BMI-specific serum Vitamin D levels of Group-1 study population (n=125)

There was no correlation between serum vitamin D level and age (r = –.05511; p = .5416; 95% CI: –0.2285, 0.1217). A significant negative linear correlation was found between vitamin D and BMI in the Pearson correlation coefficient test (r = –.2677; p = .0025; 95% CI: –0.4234, – 0.09662) (Figure 1). There was no significant correlation between serum 25(OH) D and serum iPTH (r = –.1635; p = .0685; 95%CI: –0.3296, 0.01249), and other relevant biochemical parameters (Table 4). Based on the Endocrine Society cut-off value (9), 86 (68.8%) participants had < 30ng/ml vitamin D Levels (Table 5). There was no significant difference in serum iPTH level and other relevant biochemical parameters amongst different vitamin D status groups (Table 6).

**Figure 1:**
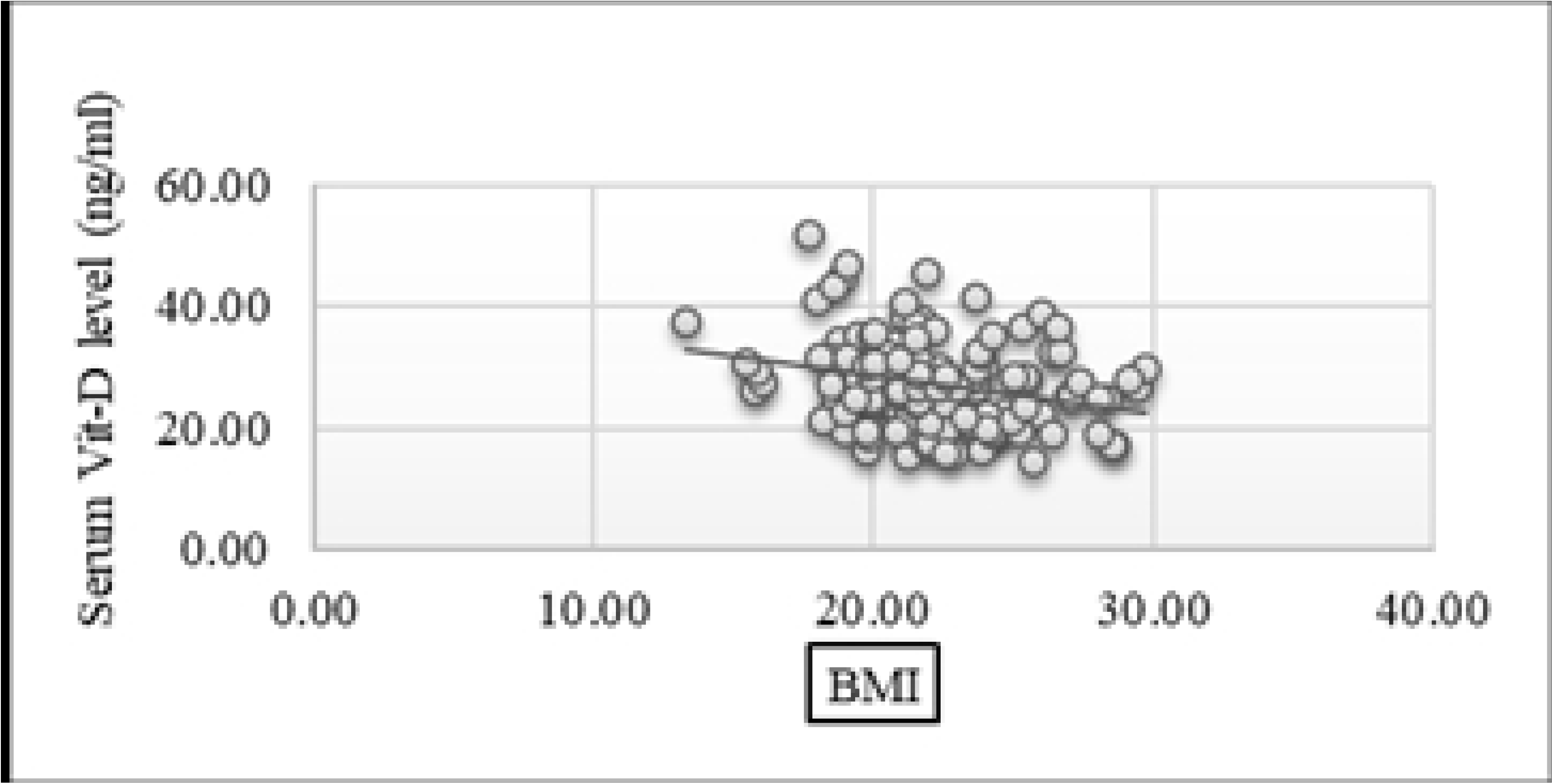
Correlation between serum 25(OH)D and BMI.

**Table 4.** Correlation between serum 25(OH)D and relevant biochemical parameters.

| Parameters | r | p-value | 95% CI for r |
| --- | --- | --- | --- |
| S. Calcium (mg/dl) | -.06803 | .4510 | -0.2408, 0.1089 |
| S. PO4 (mg/dl) | .06004 | .5060 | -0.1168, 0.2332 |
| S. Alk Phos (U/L) | -.1612 | .0725 | -0.3275, 0.01484 |
| S. iPTH (pg/ml) | -.1635 | .0685 | -0.3296, 0.01249 |

CI = confidence interval,

**Table 5:** Serum Vitamin D status of the Group-1 (coastal fishermen) study population based on the Endocrine Society cut-off value (9)

| Status | n (%) | Serum 25(OH)D<br>Mean±SD (ng/ml) |
| --- | --- | --- |
| Deficient (<20ng/ml) | 23 (18.4) | 17.96±1.69 |
| Insufficient (21ng/ml-29.9ng/ml) | 63 (50.4) | 25.52±2.53 |
| Adequate(≥30ng/ml) | 39 (31.2) | 35.90±5.16 |
| All cases | 125 | 27.36±7.27 |

**Table 6.**
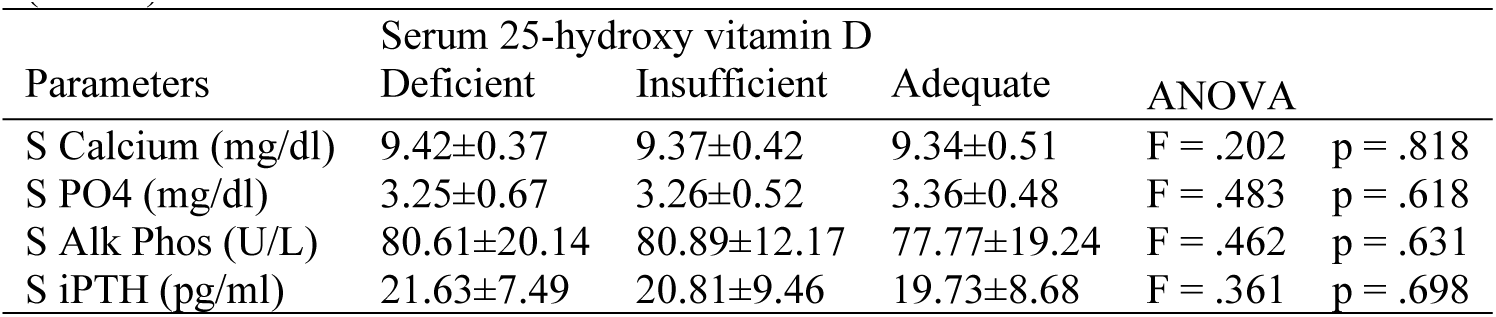
Levels of different serum biochemical markers related to vitamin D status of Groups-1 study population (n=125)

| Parameters | Serum 25-hydroxy vitamin D |  |  | ANOVA |  |
| --- | --- | --- | --- | --- | --- |
|  | Deficient | Insufficient | Adequate |  |  |
| S Calcium (mg/dl) | 9.42±0.37 | 9.37±0.42 | 9.34±0.51 | F = .202 | p = .818 |
| S PO4 (mg/dl) | 3.25±0.67 | 3.26±0.52 | 3.36±0.48 | F = .483 | p = .618 |
| S Alk Phos (U/L) | 80.61±20.14 | 80.89±12.17 | 77.77±19.24 | F = .462 | p = .631 |
| S iPTH (pg/ml) | 21.63±7.49 | 20.81±9.46 | 19.73±8.68 | F = .361 | p = .698 |

Reference interval of vitamin D of coastal fishermen (Reference population) Based on CLSI Guidelines C28-A3, 90% CI was used to calculate the reference interval of the Group-1 reference population (coastal fishermen). The calculated reference interval of serum vitamin D value by parametric, non-parametric and robust methods were 15.78-44.29, 15.88-45.27 and 15.69-44.68 ng/ml, respectively (Figure 2,Table 7). Later in the study, the vitamin D status of the urban population was determined based on the fishermen’s reference interval.

**Figure 2:**
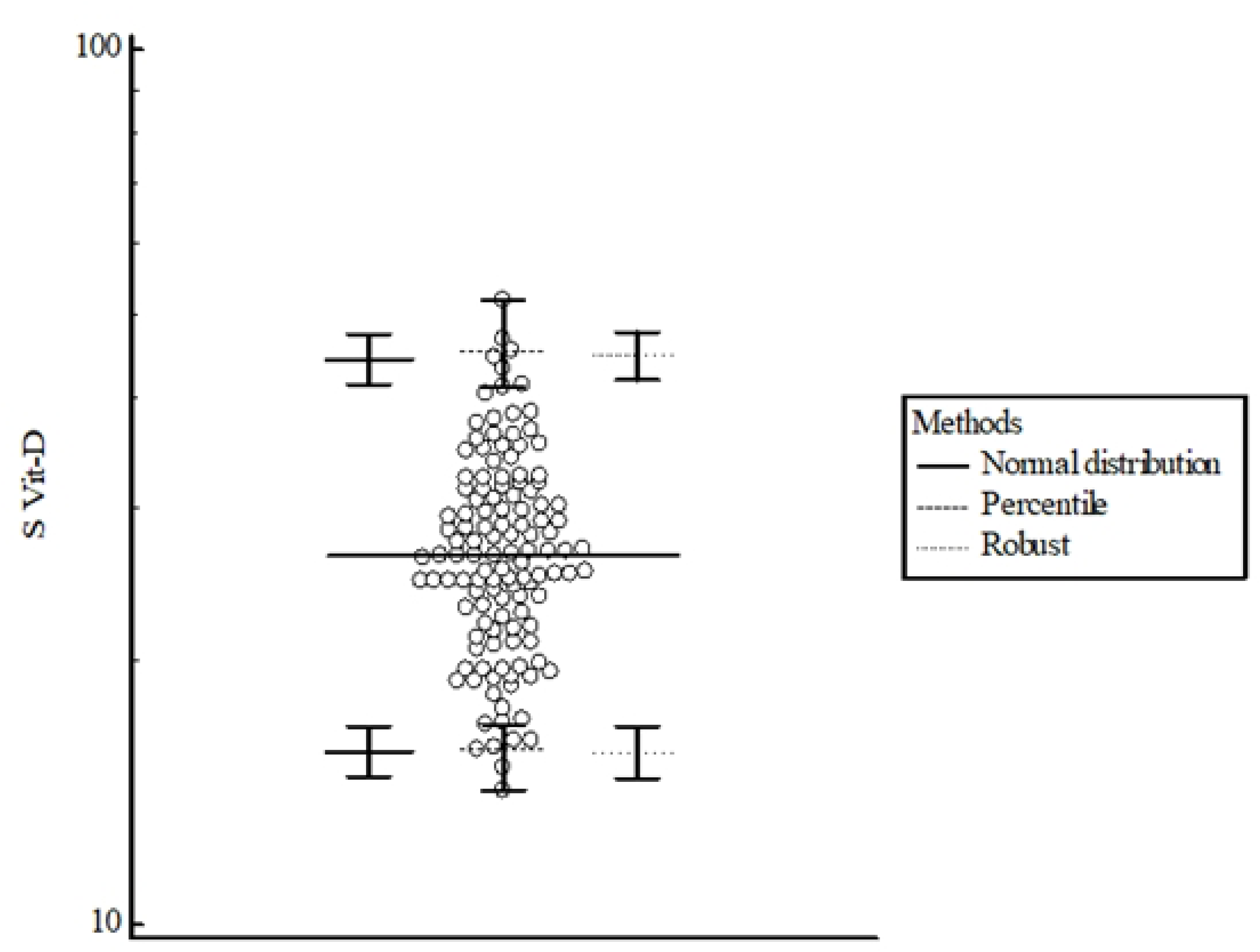
Reference interval of vitamin D. (95% Reference interval, Double-sided)

**Table 7.** Reference interval of vitamin D levels calculated by different methods using serum 25(OH)D level of Group-1 study population.

| Methods | Serum 25(OH)D (ng/ ml) |
| --- | --- |
| A. Based on Normal distribution |  |
| Lower limit (90% CI) | 15.78 (14.7581, 16.8828) |
| Upper limit (90% CI) | 44.29 (41.4114, 47.3733) |
| B. Non-parametric percentile method (CLSI C28-A3) |  |
| Lower limit (90% CI) | 15.87 (14.2600, 16.9200) |
| Upper limit (90% CI) | 45.2670 (41.1100, 51.8500) |
| C. Robust method (CLSI C28-A3) |  |
| Lower limit (90% CI<sup>a</sup>) | 15.6855 (14.6794, 16.8548) |
| Upper limit (90% CI<sup>a</sup>) | 44.6759 (41.8912, 47.5948) |

90% CI = 90% confidence interval.
<sup>a</sup>Bootstrap confidence interval (10000 iterations; random number seed: 978).

The mean vitamin D level in the urban population was 21.53±15.98 ng/ml (95% CI:19.9, 23.16), with a median of 17.18 ng/ml and a range of 3.48 to 147.32 ng/ml. There was no significant difference in vitamin D levels between male (20.16±16.18 ng/ml) and female (22.76±15.74 ng/ml) participants using parametric methods (p > .05, 95% CI: –5.86, 0.66). However, nonparametric methods indicated significantly higher levels in females (p=.008). Table 8 shows that mean serum vitamin D levels were significantly higher in older age groups (p = .001) and positively correlated with age in urban populations (r = .174, p = .001, 95% CI:0.073, 0.271; Figure 3). There were no significant differences in vitamin D levels between BMI groups (F = 0.173, p > .05).

**Figure 3:**
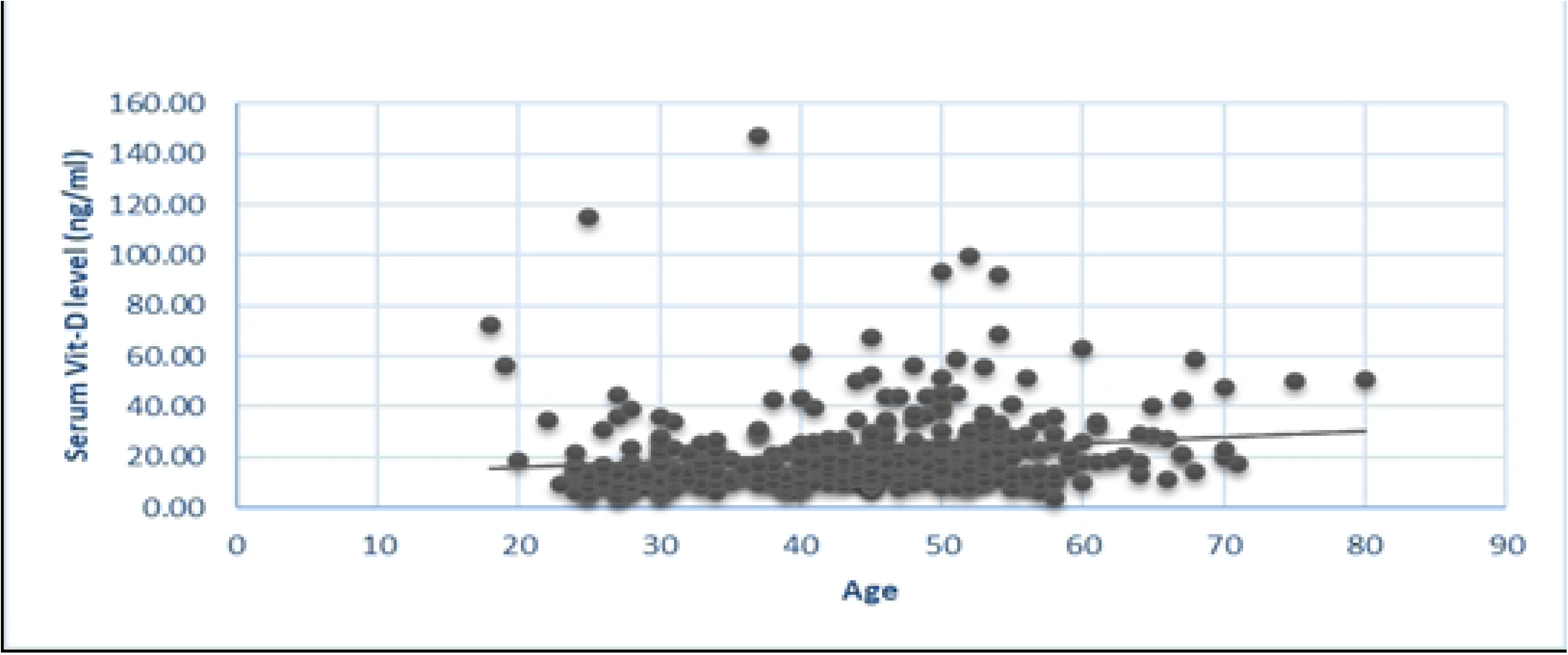
Correlation between age and vitamin D.

**Table 8.**
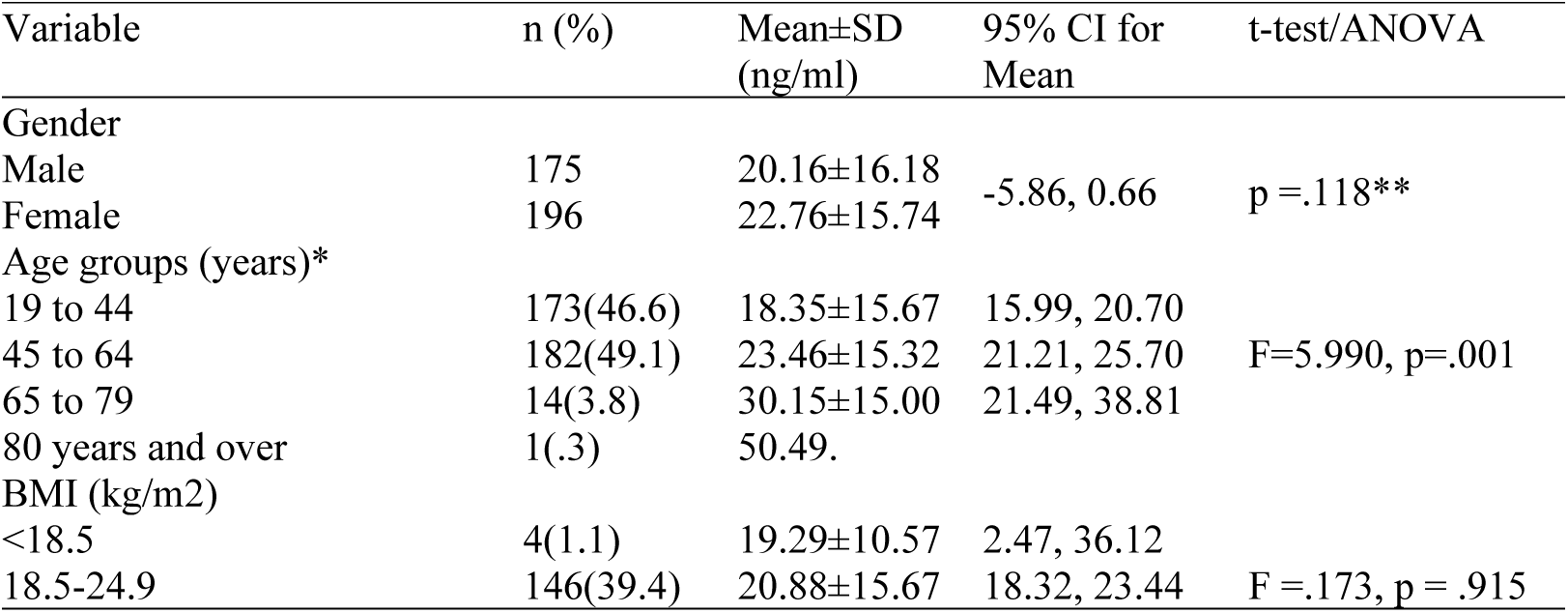
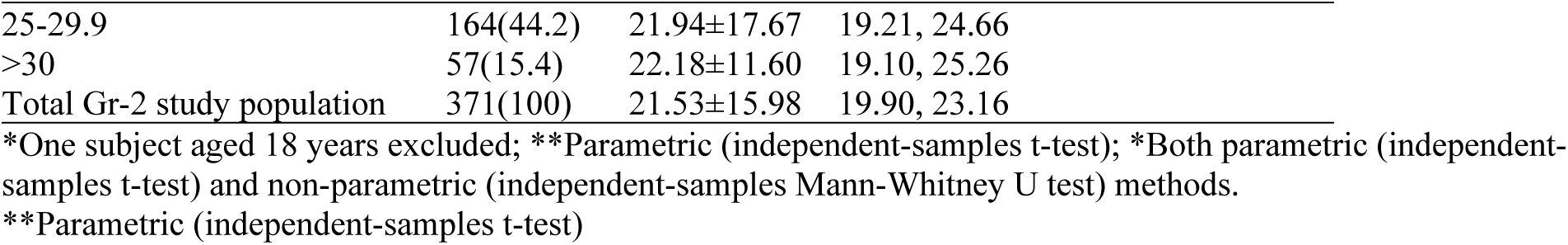
Age (MeSH classification) and BMI (WHO classification) specific serum Vitamin D levels of Group-2 study population (n=371)

| Variable | n (%) | Mean±SD<br>(ng/ml) | 95% CI for<br>Mean | t-test/ANOVA |
| --- | --- | --- | --- | --- |
| Gender |  |  |  |  |
| Male | 175 | 20.16±16.18 | -5.86, 0.66 | p =.118** |
| Female | 196 | 22.76±15.74 |  |  |
| Age groups (years)* |  |  |  |  |
| 19 to 44 | 173(46.6) | 18.35±15.67 | 15.99, 20.70 | F=5.990, p=.001 |
| 45 to 64 | 182(49.1) | 23.46±15.32 | 21.21, 25.70 |  |
| 65 to 79 | 14(3.8) | 30.15±15.00 | 21.49, 38.81 |  |
| 80 years and over | 1(.3) | 50.49. |  |  |
| BMI (kg/m2) |  |  |  |  |
| <18.5 | 4(1.1) | 19.29±10.57 | 2.47, 36.12 | F =.173, p = .915 |
| 18.5-24.9 | 146(39.4) | 20.88±15.67 | 18.32, 23.44 |  |

|  |  |  |  |
| --- | --- | --- | --- |
| 25-29.9 | 164(44.2) | 21.94±17.67 | 19.21, 24.66 |
| >30 | 57(15.4) | 22.18±11.60 | 19.10, 25.26 |
| Total Gr-2 study population | 371(100) | 21.53±15.98 | 19.90, 23.16 |

\*One subject aged 18 years excluded; \*\*Parametric (independent-samples t-test); \*Both parametric (independent-samples t-test) and non-parametric (independent-samples Mann-Whitney U test) methods.
\*\*Parametric (independent-samples t-test)

According to the Endocrine Society cut-off value (9), 83.6% of the urban population had below-normal (inadequate) vitamin D levels. However, using the fishermen’s reference intervals established in the first phase of this study, only 44.2% of the urban population were found to have below-normal vitamin D levels (Table 9). Approximately 47% of individuals classified as having inadequate vitamin D levels by the Endocrine Society’s criteria were actually within the adequate range based on the fishermen’s reference interval (**Error! Reference source not found.**). The mean serum vitamin D level was significantly lower in the urban group (21.53±15.98 ng/ml) compared to fishermen (27.36±7.27 ng/ml); 95% CI [-7.91, –3.76], p < .001.

**Table 9.** Vitamin D status of the urban people based on the Endocrine society cut-off value and fishermen’s reference interval.

| Vitamin D status of the Gr-2 people | Below normal<br>n (%) | Normal<br>n (%) | Above normal<br>n (%) |
| --- | --- | --- | --- |
| Based on ES cut-off value | 310 (83.6)<br>Deficiency 221(59.6)<br>Insufficiency 89(24.0) | 27(7.3) | 34(9.2) |
| Based on fishermen's cut-off value | 164(44.2) | 184(49.6) | 23(6.2) |

ES = Endocrine society, FM = coastal fishermen (Gr-1)

The mean serum iPTH level in the urban population was 62.80 ±31.67 pg/ml, with a median of 56.40 pg/ml, ranging from 10.20 to 227.54 pg/ml. There was a significant negative correlation between serum vitamin D and serum iPTH (r = –0.233, 95% CI: –0.3273, –0.1347; p < .001) (Figure). However, the significant differences in PTH values among different vitamin D segments indicate a nonlinear relationship between vitamin D and PTH (Table 10).

**Table 10.** Mean PTH level in different vitamin D segments based on Endocrine society cut-off value.

| Vit D status | ng/mL | n | iPTH(pg/ml)<br>Mean* | 95% CI of<br>mean |
| --- | --- | --- | --- | --- |
| Vitamin D deficiency | <20 | 221 | 66.94 | 62.68, 71.21 |
| Vitamin D insufficiency | 20 - 29.9 | 89 | 60.47 | 53.68, 67.26 |
| Vitamin D normal range | 30-40 | 27 | 59.89 | 49.89, 69.90 |
| Vitamin D above normal range | >40 | 34 | 44.33 | 35.90, 52.76 |

\* Difference of means is significant at <1% level of significance (F=5.55, p=.001)

The LOWESS (locally weighted scatter plot smoothing) method was employed to illustrate the relationship between serum iPTH and serum 25(OH)D. It was found that as vitamin D level decreased, PTH level decreased until reaching a plateau (Figure 5). This plateau, known as the PTH deflection point, indicates the lower cut-off for optimal serum vitamin D levels. Using piecewise linear regression with the Python plugin in SPSS, the visually estimated deflection point of 12.5 ng/ml was refined to 12.16 ng/ml, with a 95% confidence interval ranging from 11.04 ng/ml to 13.28 ng/ml.

**Figure 4:**
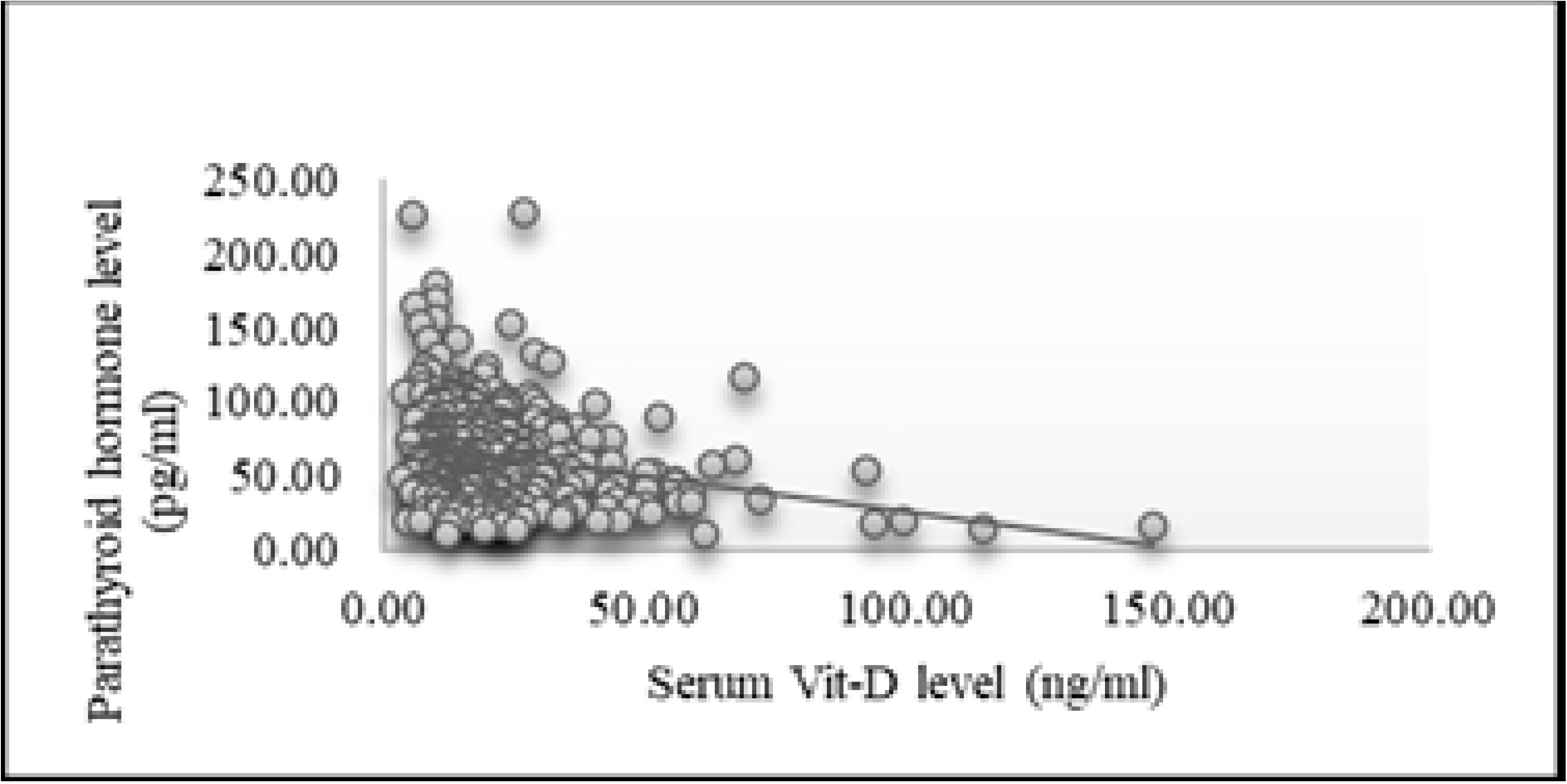
Correlation between serum vitamin D and serum iPTH in urban population.

**Figure 5:**
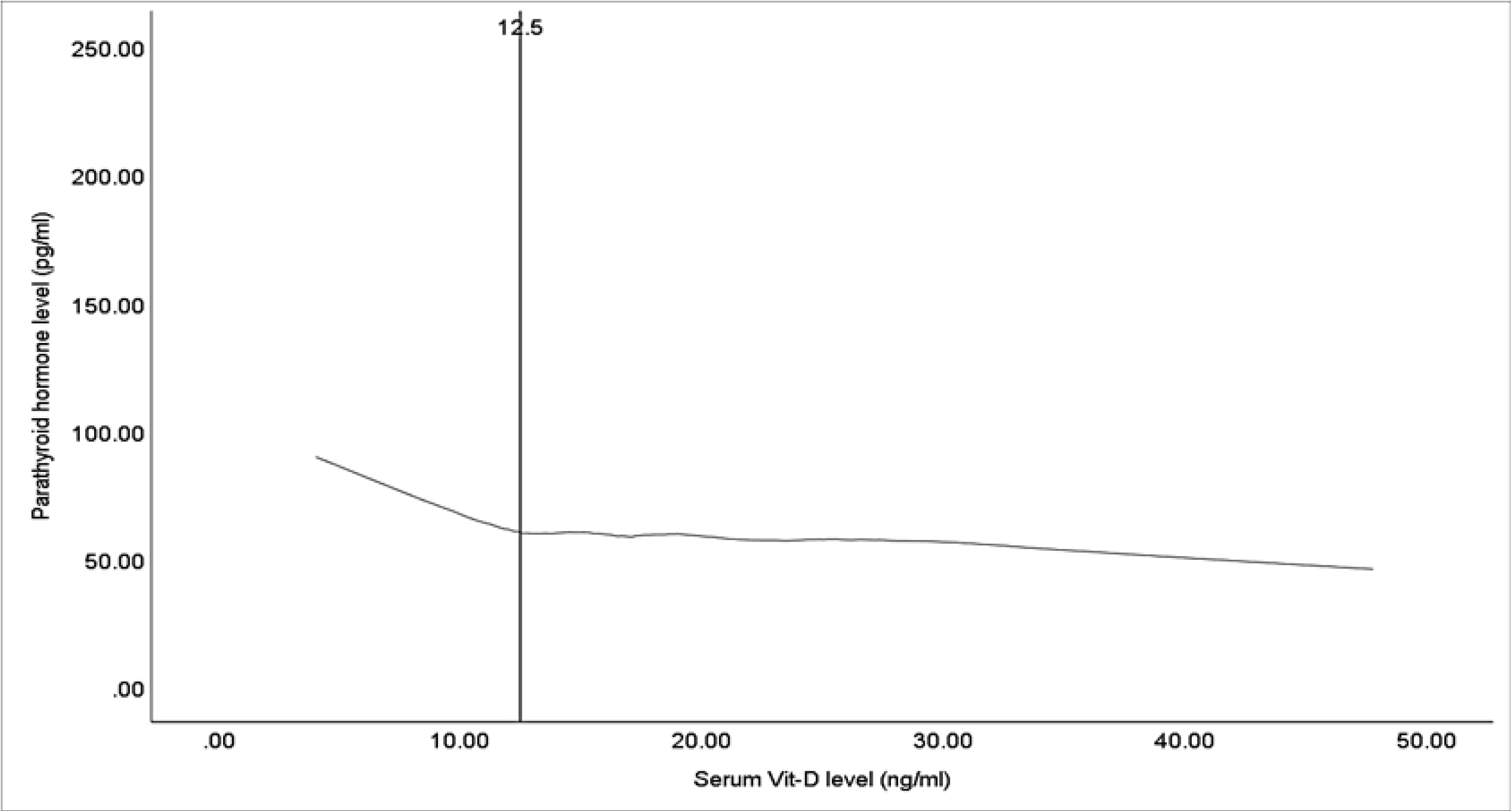
The scatterplot with LOESS fitting (50%) illustrates the relationship between vitamin D and PTH, with vitamin D (ng/ml) on the x-axis and serum iPTH (pg/ml) on the y-axis. The vertical line indicates the PTH deflection point using the raw data.

## Discussion

### Health

The study focused on the health of Group-1 (coastal fishermen), ensuring all participants were clinically and biochemically healthy. This prerequisite allowed for an accurate determination of the reference interval for vitamin D. The coastal environment, with its pollution-free air, excellent sun exposure, and consumption of marine fish, ensured vitamin D adequacy for this group. Prior studies, such as the 1958 birth cohort study in Scotland, have also linked coastal climates with higher levels of 25(OH)D due to increased sun exposure (16). The inclusion of only male participants in Group 1 to ensure adequate sun exposure might be seen as a limitation. However, other studies indicate no significant gender-specific differences in vitamin D levels (8, 17).

Overweight subjects were excluded from Group 1, yet a significant negative correlation between vitamin D and BMI was observed, consistent with previous findings (18–21). Studies consistently show an inverse relationship between BMI and vitamin D levels, highlighting the impact of body weight on vitamin D status (22–24). The lack of correlation between age and serum vitamin D levels in Group 1 suggests the overall good health of the study population. This may obscure the typical association between lower vitamin D levels and older age, often attributed to reduced outdoor activity and impaired kidney function in older adults (25).

Health status of the Group-2 participants was not as strictly controlled, with only those with acute illnesses being excluded. This approach aligns with other studies examining the correlation between PTH and vitamin D, which often include people with a broader range of health statuses (26–32).

### Vitamin D Status

In the present study, irrespective of the adequacy of sun exposure, most participants in both the fishermen and urban groups had low vitamin D levels, with means of 27.36±7.27 ng/ml and 21.53±15.98 ng/ml, respectively. This high prevalence of vitamin D inadequacy aligns with recent studies reporting widespread vitamin D insufficiency or deficiency in Bangladesh (8, 33–39). For instance, Mahmood et al. (2017) found that 100% of garment workers and 97% of agricultural and construction workers had low vitamin D levels. Garment workers, in particular, are at high risk due to limited sun exposure and poor nutritional status (40).

Within urban participants, hospital staff had lower vitamin D levels and a higher prevalence of inadequacy compared to healthy volunteers. This finding is supported by multiple studies indicating a high prevalence of vitamin D inadequacy among health personnel (41–46). The positive correlation between vitamin D and age in the urban population may be explained by higher supplement use among older individuals (43, 47, 48). Vitamin D supplementation, as shown in various studies, significantly increases serum 25(OH)D levels (43, 49).

The higher vitamin D levels in coastal fishermen compared to urban dwellers underscore the importance of sunlight exposure. Similar studies, such as those by Lee et al. (2018) and Haddad and Chyu (1971), also reported higher vitamin D levels in individuals with significant sun exposure. Despite this, around 70% of coastal fishermen in this study had low vitamin D levels according to the Endocrine Society’s cut-off, raising questions about the validity of the use of current threshold values. Previous studies in heavily sun-exposed regions in India, report similar findings of low vitamin D levels despite abundant sun exposure (50, 51).

### Reference Interval and Cut-off Point

The reference intervals for vitamin D established in this study using healthy coastal fishermen population may represent the optimal levels for the Bangladeshi population. The reasoning behind this conclusion lies in both the nature of the studied group and the limitations of previous definitions of “optimal” vitamin D status.

Historically, earlier studies that defined vitamin D thresholds were methodologically flawed. Those studies failed to adequately consider factors such as sun exposure, health, and geographic variation (52–54). The use of populations with insufficient sun exposure led to a skewed understanding of vitamin D requirements. For example, previous normative ranges (10-55 ng/mL) did not accurately capture true vitamin D deficiency, especially in groups with limited sun exposure. In contrast, populations like coastal fishermen, who have high exposure to sunlight, provide a more accurate picture of what might be the “natural” or optimal vitamin D levels for people living in sun-rich environments like Bangladesh.

In this study, a reference interval of 15.88 ng/mL (90% CI: 14.26–16.92) to 45.27 ng/mL (90% CI: 41.11–51.85) was determined using non-parametric methods aligning with the Clinical and Laboratory Standards Institute (CLSI) Guidelines(55). The non-parametric approach is appropriate given the asymmetry of the data, providing a robust understanding of the distribution in the fishermen population. The upper limit of 45.27 ng/mL aligns with the commonly accepted upper limit proposed by the Endocrine Society, suggesting the upper threshold of vitamin D status is consistent across various populations.

However, the lower limit of 15.88 ng/mL stands below the traditionally accepted threshold levels (often cited as 20-30 ng/mL). This finding is supported by evidence from other population where lower cut-off values for vitamin D sufficiency have been proposed. For instance, studies from South Korea (56), India (30), China (28), and Greece (29) have consistently shown lower thresholds of 13-20 ng/mL. The range in this study is closer to those findings, implying that vitamin D requirements are likely influenced by both environmental and genetic factors, and that a universal cut-off point may not be applicable globally.

Moreover, PTH dynamics provide additional support for these lower thresholds. Vitamin D inadequacy is often marked by increased PTH levels, which serve as a surrogate marker for deficiency. However, in this study, no significant correlation between vitamin D and PTH levels was found in the coastal fishermen, unlike in the urban population, where a deflection point in PTH occurred at levels far below the Endocrine Society’s recommended 30 ng/mL. Studies involving other population have noted similar findings: PTH rises as vitamin D level drops below 8-21.1 ng/mL (26, 28–30, 32, 57–60). These studies suggest that a threshold for PTH deflection, and thus vitamin D sufficiency, may indeed be lower than previously thought. The absence of PTH deflection in coastal fishermen may indicate the overall vitamin D adequacy of this population.

Additional support in favour of our low reference interval and lower cut-off values for vitamin D sufficiency comes from recent randomized controlled trials (RCTs) and cohort studies. For instance, post hoc analyses from trials like VITAL and ViDA revealed that vitamin D supplementation benefited participants only when baseline levels were below 12-15 ng/mL (61–63). Martineau et al. (2017) noted that vitamin D supplementation was most effective in preventing respiratory infections in participants with baseline levels below 10 ng/mL (64). These findings argue against higher supplementation thresholds and support a more moderate cut-off for deficiency. Furthermore, based on epidemiological evidence, Manson et al. (2016) and the Institute of Medicine (IOM), supports a deficiency threshold closer to 12.5 ng/mL(63, 65). Studies in pregnancy and diabetes prevention also point toward 15 ng/mL as a sufficient level for avoiding adverse outcomes (31, 66).

Based on this evidence, it is logical to conclude that the reference interval identified in the current study (15.88–45.27 ng/mL) could reflect the optimal range for the Bangladeshi population. The high sun exposure in coastal fishermen makes this population an ideal reference group, better reflecting the biological requirements of individuals in sun-rich environments like Bangladesh. Moreover, the lower limit of this range is consistent with thresholds suggested by contemporary studies that challenge higher vitamin D cut-off points. Thus, considering the surrogate evidence, PTH dynamics, and recent trial data, this study provides a more region-specific, evidence-backed approach in determining optimal vitamin D levels.

### Strengths and Limitations of this Study

- The study employed a cross-sectional design, sampling both coastal fishermen and urban residents to establish the reference interval of vitamin D and determine the lower cutoff value.
- Reference intervals were established following the Clinical and Laboratory Standards Institute (CLSI) Guidelines C28-A3 for precise comparison across populations, and the lower cutoff value was identified using the PTH deflection point.
- Data collection included structured questionnaires, anthropometric measurements, and standardized biochemical testing to ensure accuracy and reliability.
- The study did not employ strict random sampling, which may introduce selection bias and limit generalizability.
- The absence of a gold-standard LC-MS/MS vitamin D assay may affect the precision of biochemical measurements.

### Conclusion and Recommendations

Vitamin D, an essential micronutrient, is endogenously produced when exposed to sunlight. Despite the global prevalence of vitamin D deficiency/insufficiency, landmark trials have not justified routine vitamin D supplementation for the general population. The optimal vitamin D level remains controversial and may vary depending on population and geography. Bangladesh, with its ample sunlight, still faces widespread vitamin D inadequacy according to current reference values. This study’s findings on healthy coastal fishermen provide critical insights into the optimal serum vitamin D levels for Bangladeshis. Additionally, the study has defined the lower normal limit of vitamin D sufficiency by determining the serum iPTH deflection point, suggesting a lower optimal range than currently used.

Thus, the findings of the present study would help in guiding clinicians and policymakers in developing appropriate treatment and supplementation guidelines for Bangladeshi population.

## Data Availability Statement

The dataset supporting the conclusions of this study is available in the Zenodo repository, DOI: <u>10.5281/zenodo.13950605</u>. Additional materials, such as the technical appendix and statistical code, can also be accessed at this DOI.

## Author Contributions

Professor **Wasim Md Mohosin Ul Haque** made substantial contributions as the primary researcher, leading the data collection, analysis, and drafting of the manuscript. Professors **Jalaluddin Ashraful Haq**, **Md. Faruque Pathan**, and **Mohammed Abu Sayeed** contributed equally to the conception, design, interpretation of the data, and critical revision of the manuscript for important intellectual content. All authors provided final approval of the version to be published and agree to be accountable for all aspects of the work, ensuring that any questions related to the accuracy or integrity of any part are appropriately investigated and resolved.

## Corresponding author

Professor Wasim Md Mohosin Ul Haque E-mail ID, Mobile number: +8801915472750

## Co-authors

Professors Jalaluddin Ashraful Haq

Professors Md. Faruque Pathan

Professors Mohammed Abu Sayeed

